# Evaluation of the current therapeutic approaches for COVID-19: a meta-analysis

**DOI:** 10.1101/2020.08.24.20180638

**Authors:** Zeinab Abdelrahman, Qian Liu, Shanmei Jiang, Mengyuan Li, Yue Zhang, Xiaosheng Wang

**Affiliations:** Biomedical Informatics Research Lab, School of Basic Medicine and Clinical Pharmacy, China Pharmaceutical University, Nanjing 211198, China.; Big Data Research Institute, China Pharmaceutical University, Nanjing 211198, China; Pinghu Hospital of Shenzhen University, Shenzhen 440307, China; Futian Hospital for Rheumatic Diseases, Shenzhen 518000, China; Department of Rheumatology and Immunology, The First Clinical College of Harbin Medical University, Harbin 150001, China

**Keywords:** COVID-19 drug treatment, COVID-19, Antiviral Agents, Interferons, Hydroxychloroquine, tocilizumab

## Abstract

**Background and rationale:** Limited data on the efficacy and safety of currently applied COVID-19 therapeutics and their impact on COVID-19 outcomes have raised additional concern.

**Aim and Methods:** We estimated the impact of the current treatments on the efficacy and safety of COVID-19 by a meta-analysis. The comprehensive search included studies reporting clinical features and treatment strategies published from January 21, 2020, to May 15, 2020.

**Results:** We included 52 studies that involved 13,966 COVID-19 patients. We found that the most prevalent treatments were antivirals (proportion: 0.74, 95% CI^1^: [0.65, 0.83]) and antibiotics (proportion: 0.73, 95% CI: [0.62, 0.83]). The COVID-19 severity increased among patients taking glucocorticoids (risk ratio (RR)^2^ = 1.71, 95% CI: [1.06, 2.76]) or immunoglobulins (RR = 3.83, 95% CI: [1.27, 11.53]), and renal replacement therapy (RRT) and glucocorticoids increased the length of ICU stay (RRT^3^: RR = 11.89, 95% CI: [3.26, 43.39]; glucocorticoids: RR = 3.10, 95% CI: [1.52, 6.29]). The COVID-19 severity and mortality increased among patients taking tocilizumab (severity: F = 25.53, *P* = 0.02; mortality: F^4^ = 19.37, *P* = 0.02). The most effective treatment was the combination of arbidol with lopinavir/ritonavir compared with placebo (mean difference = 0.5, 95% CI [-0.60, 1.66]), and the safest combination was remdesivir and lopinavir/ritonavir (RR = 0.78, 95% CI [0.32, 1.91]).

**Conclusion:** glucocorticoids, immunoglobulins, RRT, and tocilizumab might worsen COVID-19 outcomes, and themost effective and safest treatment strategy for COVID-19 is the combination of different antivirals.

## 1. Introduction

Since the outbreak of the coronavirus disease 2019 (COVID-19) caused by severe acute respiratory syndrome coronavirus 2 (SARS-CoV-2) in December 2019, more than 14 million cases and 613,879 deaths have been reported as of July 22, 2020, in the world^[1]^. Compared to other beta coronaviruses that have caused epidemics over the last two decades, including severe acute respiratory syndrome coronavirus (SARS-CoV)^[2]^ and Middle East respiratory syndrome coronavirus (MERS-CoV)^[3]^, SARS-CoV-2 exhibits higher infectivity while lower fatality that makes it more destructive^[4]^. The rapid development of effective treatment approaches for COVID-19 is urgently needed since there is no specific therapy or vaccine for COVID-19 currently. Previous experiences from SARS and MERS treatment suggest that several interventions, including antivirals, such as lopinavir/ritonavir and umifenovir, glucocorticoids, interferons, ribavirin, besides newly introduced drugs such as chloroquine or its derivative hydroxychloroquine, and convalescent plasma, may improve clinical outcomes in COVID-19 patients, whereas the related data are not conclusive. Since the evidence of the effectiveness of these treatments remains lacking, we conducted a meta-analysis of the therapeutic approaches for COVID-19. This meta-analysis aimed to identify favorable and unfavorable treatment strategies for COVID-19 by comparing different treatment approaches using machine learning models.

First, we conducted a proportional meta-analysis to summarize the pooled effect of the weighted proportion of each treatment. Next, we identified the impact of each treatment on the severity and mortality of COVID-19. Finally, we conducted the network meta-analysis to, directly and indirectly, compare different treatment strategies. We summarized available randomized and non-randomized clinical trials of several treatment strategies and provided point estimates and their 95% confidence intervals (CIs) for the associations between these treatment strategies and given endpoints.

## 2. Methods

### 2.1 Search strategy and selection criteria

We searched for publications between January 21, 2020, and May 15, 2020, in databases of PubMed, EMBASE, CNKI, Wanfang, Cochrane library, http://ClinicalTrails.gov, Scopus, Lancet, N Engl J Med and JAMA platforms, and Web of Science. We used the EndNote X9.0 software to exclude duplicate or irrelevant articles. We used the search term “2019 novel coronavirus, COVID-19 and SARS-CoV-2” AND “treatment, clinical characteristics, epidemiological characteristics, clinical trials, cohort studies, observational studies, case series” without language and age restriction. To identify missed studies, we checked the reference list for each selected paper. Our meta-analysis included publications involving the following information: clinical features, laboratory findings, and treatment approaches for COVID-19 patients with clinically defined severity or mortality. We excluded the following studies: duplicate publications, preprints, reviews, case reports, family-based studies, unrelated titles or abstracts, studies not involving clinical features, laboratory findings, treatment approaches, and animal or in vitro studies. The literature search steps followed the transparent reporting of systematic reviews and meta-analyses “PRISMA” (Figure 1, Supplemental Content 1).

**Figure 1.**
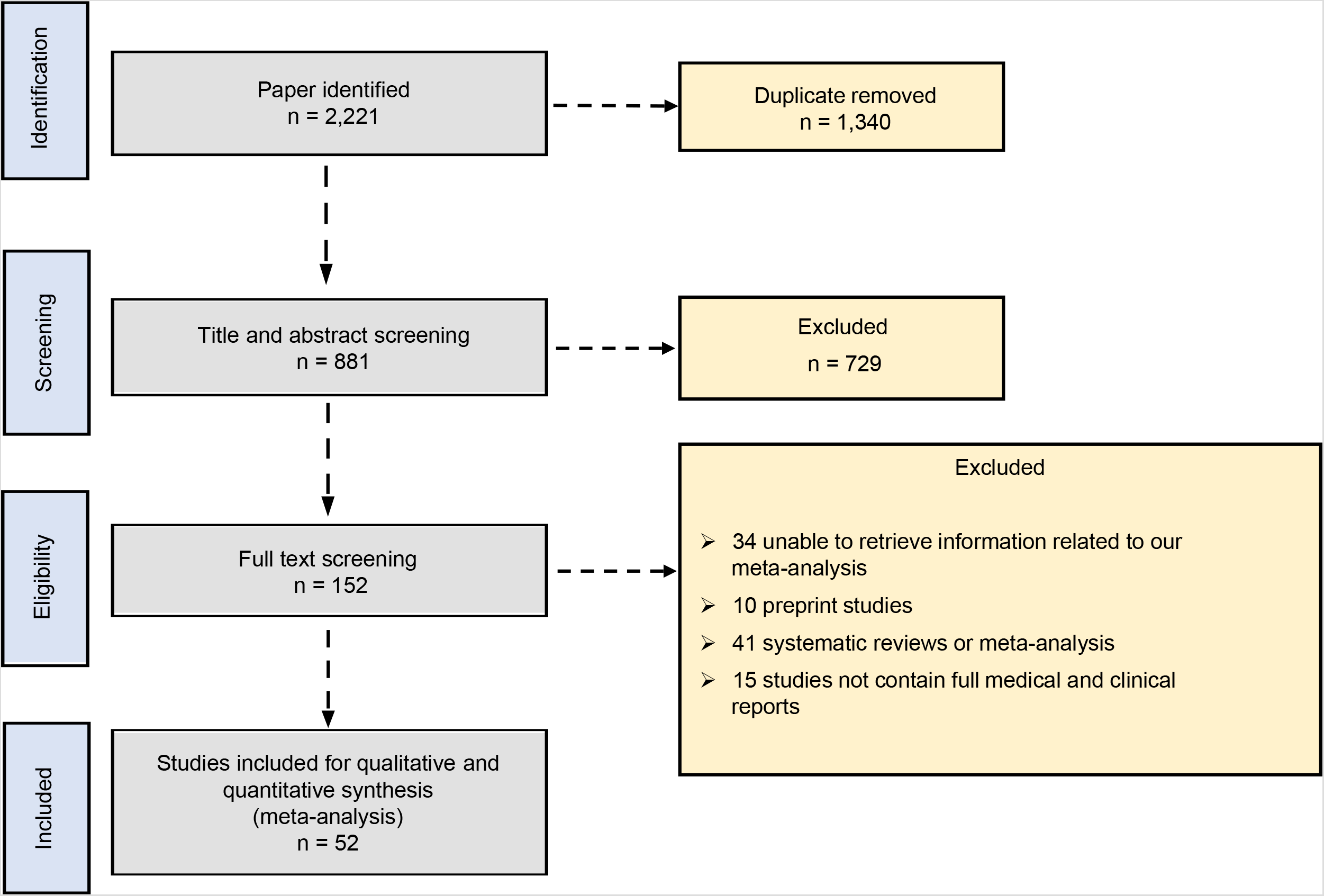
PRISMA 2009 flow diagram.

### 2.2 Data extraction

Two investigators (QL and SJ) performed a literature search and data extraction, and another investigator (ZA) resolved the disagreements. We extracted the following variables: author, date, age, gender, number of participants in different groups for comparisons, including severe versus non-severe, ICU versus non-ICU, death versus survival, and deterioration versus discharge. The extracted data included publication date, country, study design, number of enrolled subjects, data collection method, baseline characteristics before treatment, diagnostic method, population, treatment details, time from admission to starting treatment, and outcomes in patients.

### 2.3 Data analyses

We conducted a proportional meta-analysis to account for the weighted average proportion of each treatment. We also conducted the subgroup meta-analysis to identify the impact of each treatment on the disease severity, such as the utility of antibiotics or antivirals in severe versus non-severe, ICU versus non-ICU, death versus survival, and deterioration versus discharge patients. Besides, we conducted the network meta-analysis to account for the relationship between clinical outcomes and borrowed strength across studies. We defined the disease severity as a dichotomous variable. We normalized the data by double-arcsine- or logit-transformation and confirmed their normal distribution by the Shapiro-Wilk test before meta-analyses. We assessed statistical heterogeneity using the *I^2^* statistic and Cochran’s Q test with a threshold of *I^2^* ≤ 50% or *P*≤ 0.05. We adopted a random-effect model in terms of statistical heterogeneity. We performed the meta-analysis using R software and Cochrane software REVMAN 5^[5]^. For the proportional meta-analysis, we used the R package *“metafor”* and *“meta”*^[6]^ based on the restricted maximum likelihood method. We estimated the summary proportional effect size as the weighted average of the observed effect sizes in individual studies.

For subgroup meta-analysis, we applied random models and mixed-effect meta-regression for different outcomes. The subgroups were defined by different severity definition terms in the studies. We used the random-effect model in the subgroup meta-analysis, and in the meta-regression, we used the mixed-effect model with the risk ratio (RR) as the estimated effect size to evaluate the impact of different drugs on the disease severity. We also conducted the network meta-analysis of 10 different drugs in 13 studies to estimate if one intervention is more or less effective than another intervention for COVID-19 patients. In the network meta-analysis, we used a random-effect model with the RR as the size effect and estimated the network inconsistency.

### 2.4 Study selection and risk of bias assessment

We assessed the risk of bias eligible observational studies, such as cross-sectional, cohort studies, and case series, following the Strengthening the Reporting of Observational Studies in Epidemiology (STROBE) reporting guidelines^[7]^. We assessed the risk of bias in randomized control trials using the Cochrane risk-of-bias tool for randomized trials (ROB-2)^[8]^. We conducted both risks of bias evaluations by the two investigators (QL and SJ) independently, each assigned an overall risk of bias to each eligible study, and consulted a third reviewer (ZA) if they disagreed. We summarized the results in Supplemental Content 6.

## 3. Results

We initially identified 2,221 articles based on our search criteria, most of which were irrelevant to our research objective or without published results (Figure 1). Finally, we included 52 articles ^[5, 6, 7, 8, 9, 10, 11, 12, 13, 14, 15, 16, 17, 18, 19, 20, 21, 22, 23, 24, 25, 26, 27, 28, 29, 30, 31, 32, 33, 34, 35, 36, 37, 38, 39, 40, 41, 42, 43, 44, 45, 46, 47, 48, 49, 50, 51, 52, 53, 54, 55, 56, 57, 58, 59, 60]^ in our meta-analysis, which involved a total of 13,966 subjects. A summary of these studies is presented in Supplemental Content 2.

### 3.1 Proportional meta-analysis

We performed proportional meta-analyses to estimate one-dimensional binomial (weighted) average proportions. We obtained the average of the proportions of multiple studies weighted by the inverse of their sampling variances using the random-effect model. We defined eight groups of treatments commonly used against COVID-19, as shown in Supplemental Content 3. The sample sizes in these studies ranged from 9 to 5,700, with a total number of 13,966. The treatment categories included antibiotics, antivirals, glucocorticoids, chloroquine or hydroxychloroquine, immunoglobulin, interferons, tocilizumab, and renal replacement therapy (RRT). The random-effect model estimated the prevalence of each treatment and found that the highest prevalence were antivirals (proportion: 0.74, 95% CI: [0.65, 0.83]) and antibiotics (proportion: 0.73, 95% CI: [0.62, 0.83]) (Figure 2). We showed the results of proportional meta-analysis heterogeneity in Supplemental Content 4.

**Figure 2.**
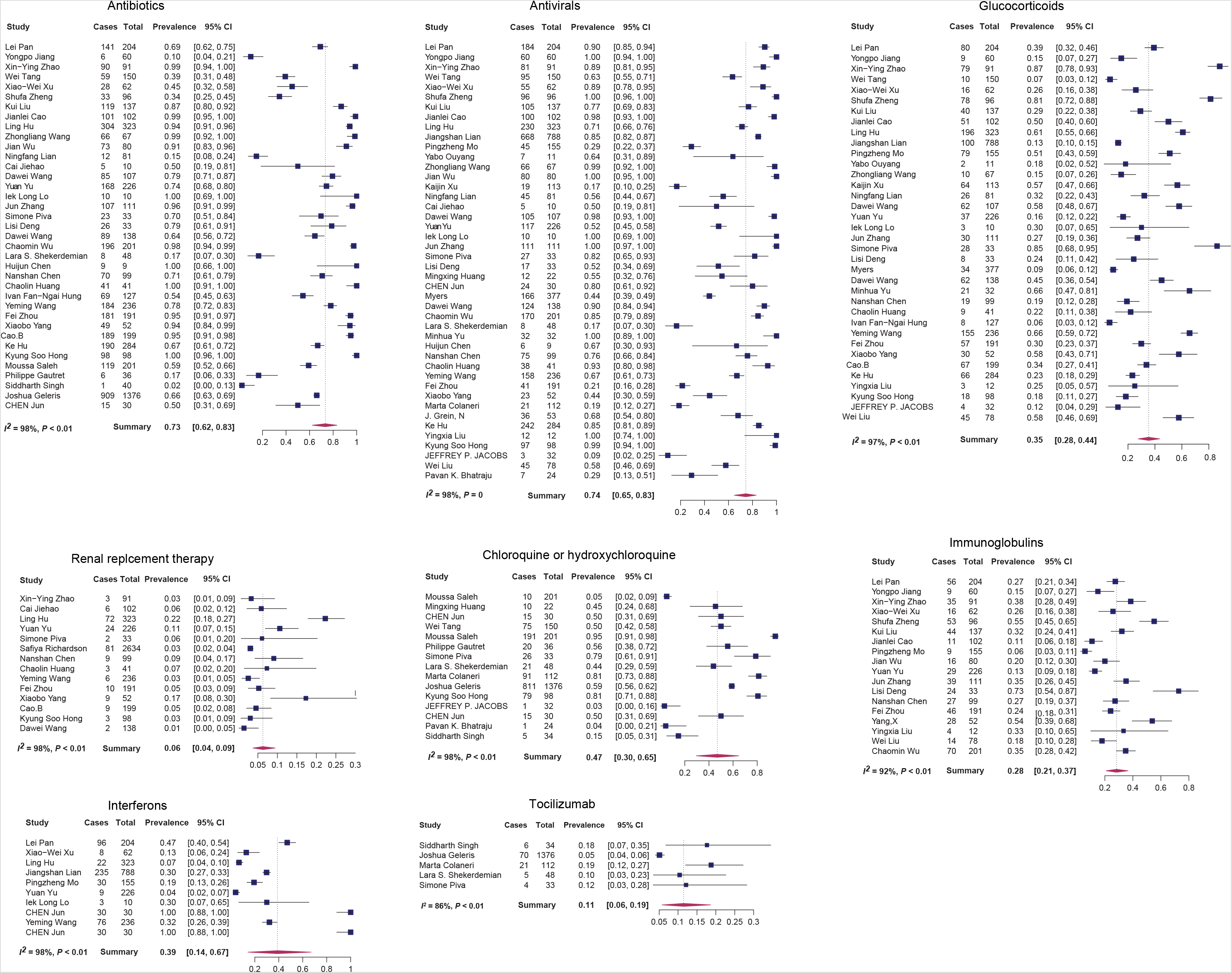
Proportional meta-analysis forest plots of the eight treatment strategies for COVID-19 by the random-effect model. Cases, number of patients taking the drug; Total, the total number of patients enrolled in the study.

To estimate the association between treatment and disease severity, we performed a subgroup meta-analysis or meta-regression to quantify each drug’s effect on the severity of COVID-19. This analysis would elucidate whether a treatment can reduce COVID-19 risk or not. The severity terms included “severe” versus “non-severe,” “death” versus “survival,” “deterioration,” versus “discharge,” and “ICU” versus “non-ICU.” We performed a subgroup meta-analysis on 42 studies. We also conducted meta-regression for chloroquine or hydroxychloroquine and tocilizumab for which we had no sufficient data to perform subgroup meta-analysis. We used the general term “relative risk (RR)” to uniformly refer to the different severity terms. The results showed that the risk of COVID-19 severity increased among patients taking glucocorticoids (RR = 1.71, 95% CI: [1.06, 2.76]) or immunoglobulins (RR = 3.83, 95% CI: [1.27, 11.53]), and RRT and glucocorticoids increased the length in ICU among COVID-19 patients (RRT: RR = 11.89, 95% CI: [3.26, 43.39]; glucocorticoids: RR = 3.10, 95% CI: [1.52, 6.29]). In addition, meta-regression analysis showed that the COVID-19 severity and mortality increased among patients taking tocilizumab (severity: F = 25.53, *P* = 0.02; mortality: F = 19.37, *P* = 0.02), but there were no significant differences among patients taking chloroquine or hydroxychloroquine. The pooled RR of eight treatments indicated that the use of glucocorticoids, immunoglobulin, tocilizumab, and RRT were likely to increase COVID-19 progression and severity (Table 1 and Supplemental Content 7 and 8).

**Table 1.** Summary of the impact of the eight treatments on COVID-19 severity.

| Treatment | Pooled effect size | P-value | Method | Heterogeneity $I^2$ (%) |
| --- | --- | --- | --- | --- |
| Glucocorticoids | Risk ratio (RR) = 2.01,<br>95% CI: [1.48, 2.74] | < 0.0001 | Subgroup meta-analysis <sup>c</sup> | 87 |
| Antibiotics | RR = 1.03,<br>95% CI: [0.99, 1.08] | 0.16 | Subgroup meta-analysis | 45 |
| Antivirals | RR = 1.00,<br>95% CI: [0.99, 1.01] | 0.93 | Subgroup meta-analysis | 0 |
| Immunoglobulins | RR = 3.09,<br>95% CI: [1.63, 5.87] | 0.0005 | Subgroup meta-analysis | 82 |
| Renal replacement therapy | RR = 10.95,<br>95% CI: [0.46, 258.94] | 0.14 | Subgroup meta-analysis | 99 |
| Interferons | RR = 1.12,<br>95% CI: [0.54, 2.30] | 0.77 | Subgroup meta-analysis | 48 |
| Chloroquine or hydroxychloroquine <sup>a</sup> | F = 0.033 | 0.85 | Meta-regression <sup>d</sup> | 97.27 |
| Chloroquine or hydroxychloroquine <sup>b</sup> | F = 0.03 | 0.87 | Meta-regression | 97.27 |
| Tocilizumab <sup>a</sup> | F = 25.53 | 0.02 | Meta-regression | 26.47 |
| Tocilizumab <sup>b</sup> | F = 19.37 | 0.02 | Meta-regression | 26.47 |

note:
<sup>a</sup> The moderator used in the meta-regression is the number of patients in ICU.
<sup>b</sup> The moderator used in the meta-regression is the number of patients died.
<sup>c</sup> Subgroup meta-analysis of severe versus non-severe, ICU versus non-ICU, death versus survival, and deterioration versus discharge by the random-effect model.
<sup>d</sup> Meta-regression of two moderators (the number of patients in ICU and the number of patients died) by the mixed-effect model.

To determine if there are outliers that could influence our analysis results, we performed leave-one-out (LOO) analyses. In brief, each study was removed once, and the summary proportion was re-estimated based on the remaining studies. Studies with a statistically significant impact on the fitted model were removed as outliers, and the model was re-fitted again. Among the eight models, we found that only in RRT, interferon, and tocilizumab models, the studies^[54, 32, 28]^had a significant impact, with Z values of 3.5, 4.5, and –5.2, respectively, and the heterogeneity reduction to 75.9%, 96.8%, and 88%, respectively (Supplemental Content 9 and 10). Finally, we assessed the potential publication bias in each treatment model using a funnel plot of the mixed-effect meta-regression model and the Egger’s regression test with the standard error as a predictor. We observed that the funnel plots were roughly symmetrical and that the Egger’s test was significant in four treatment models (antibiotics, antivirals, immunoglobulin, and RRT), indicating the clear evidence of publication bias (Supplemental Content 11).

### 3.2 Network meta-analysis

To compare the effect of different anti-COVID-19 treatment approaches, we performed network meta-analysis. We included only 13 studies ^[38, 32, 26, 24, 22, 23, 28, 60, 30, 27, 21, 29, 25]^ in the analysis because of network inconsistency (Supplemental Content 5).

First, we used the frequentist network model to define the effect size of each comparison as the mean difference (MD) and the source of network heterogeneity as the Q value. We observed the highest Q value (Q = 21.02) in the study by Tang et al. ^[23]^, and its network inconsistency was as high as 93.7%. Next, we estimated the effect of the treatments relative to the placebo, which was supplemental oxygen, noninvasive and invasive ventilation, antibiotics, vasopressor support, RRT, or extracorporeal membrane oxygenation (ECMO) (Figure 3). Many studies^[38, 26, 22, 30, 21, 60]^ have performed a comparison between lopinavir/ritonavir or remdesvir and placebo. The best improvement in the effect was the combination of arbidol with lopinavir or ritonavir (MD = 0.5, 95% CI [-0.60, 1.66]), and the next one was hydroxychloroquine (MD = 0.3, 95% CI [-0.15, 0.68]) when compared to placebo. We also estimated the percentage of direct and indirect evidence used for each comparison, which may contribute to the assumption of consistency underlying the network meta-analysis model. Nevertheless, this estimate can evaluate the extent to which the comparisons were inferred by direct or indirect evidence. We produced a matrix for the effect sizes of all possible treatment combinations and found that the combination of remdesivir and lopinavir/ritonavir had the lowest RR (RR = 0.78, 95%CI [0.32, 1.91]) (Figure 4).

**Figure 3.**
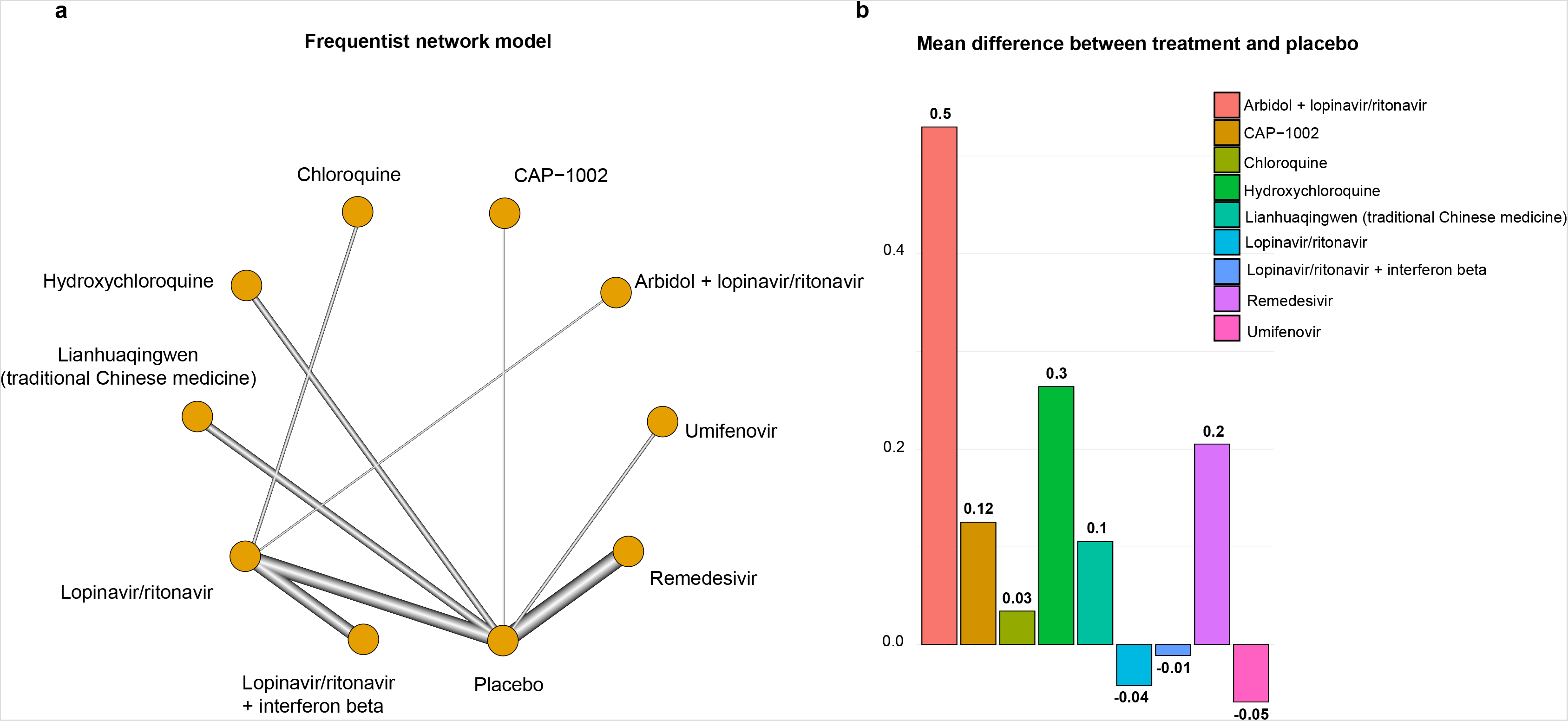
a) Network frequentist model of 10 treatment approaches for COVID-19 and b) the mean difference between each treatment and placebo. The edge thickness is proportional to the comparison frequency of both treatments, represented by the nodes the edge connects. These results were obtained by the random-effect model.

**Figure 4.**
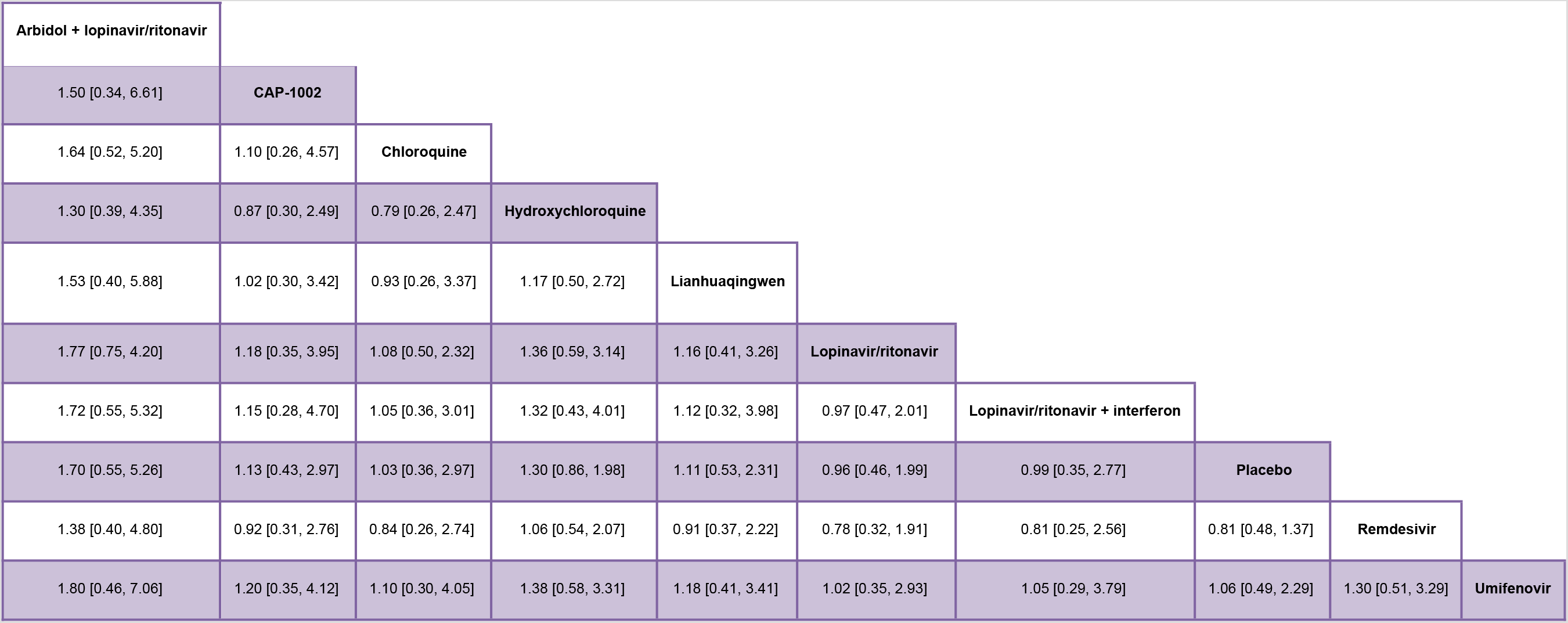
The matrix for the effect sizes of all possible treatment combinations. The risk ratios and their 95% confidence intervals are shown generated by the random-effect model.

To determine which COVID-19 treatment is the most effective, we ranked the treatments based on the P score in the frequentist model (Supplemental Content 12). We observed that arbidol plus lopinavir/ritonavir had the highest P score of 0.9995, and lopinavir/ritonavir plus interferon had the second-highest P score of 0.851, followed by chloroquine (P score = 0.579). These results indicate that the antiviral treatment has the best effect against COVID-19 and that chloroquine and its derivative are also effective.

Assessing the publication bias of a network meta-analysis in its aggregated form is problematic^[61]^. Thus, we conducted an analysis called “comparison-adjusted funnel plot” to assess the possible publication bias under an a priori hypothesis that specific mechanisms may cause the publication bias. We did not observe the funnel asymmetry (Egger’s Test, *P* = 0.38) (Supplemental Content 13).

## 4. Discussion

Currently, most of the COVID-19 treatments are symptomatic treatments, and oxygen therapy is the first step in addressing respiratory impairment. Accumulating knowledge on the pathophysiology of lung damage provides clinicians the strategies for dealing with respiratory failure caused by COVID-19^[62]^. Several treatment attempts have become an essential part of COVID-19 treatment and management protocol, such as antibiotics, antivirals, glucocorticoids, interferons, RRT, chloroquine or hydroxychloroquine, and tocilizumab, whereas there were no recommendations or rationale for using them. Earlier, 58% of COVID-19 patients in Wuhan were treated with antibiotics. Later on, the WHO recommended using empiric antibiotics to cover bacterial superinfections of COVID-19 patients^[63]^. The rationale for using glucocorticoids is still controversial, although the most common reason for using steroids is to mitigate the destructive inflammatory response in severe COVID-19 patients^[64]^. Several mechanisms have proposed that chloroquine or hydroxychloroquine could be effective against SARS-CoV-2^[65]^. However, the clinical evidence regarding the effectiveness of these treatments remains lacking. Our meta-analysis identified and summarized eight therapeutic approaches commonly used in COVID-19 treatment, including antibiotics, antivirals, immunoglobulin, glucocorticoids, interferons, chloroquine, or hydroxychloroquine, RRT, and tocilizumab. First, we found that the use of antibiotics, antivirals, and chloroquine or hydroxychloroquine was the most prevalent. Second, we found that the use of glucocorticoids, immunoglobulin, RRT, and tocilizumab was likely to promote the disease severity and deterioration among COVID-19 patients. Third, we found that the use of a combination of arbidol plus lopinavir/ritonavir had the best effect on COVID-19 patients relative to placebo, while the use of umifenovir had the worst effect. Finally, we confirmed that antivirals, especially lopinavir/ritonavir, were superior to the other anti-COVID-19 treatments.

This meta-analysis has several limitations. First, we may not include some relevant studies due to certain restrictions in searching for some databases. Second, the quality of evidence is limited by data primarily derived from retrospective analyses, which included heterogeneous data reporting and study design. Third, the confidence of the results from this meta-analysis is limited by insufficient data of the randomized control trials.

In conclusion, glucocorticoids, immunoglobulins, RRT, and tocilizumab may worsen outcomes in COVID-19. The most effective and safest treatment strategy for COVID-19 is the combination of different antivirals, supporting the use of antivirals as the basic treatment of COVID-19 with no association with disease progression and mortality. Our data are potentially valuable for clinical treatment and management of COVID-19 patients.

## Data Availability

all the data extracted in this manuscript are available online and all have been cited in the manuscript.

## Author contribution

**Zeinab Abdelrahman:** Conceptualization, Methodology, Software, Investigation, Formal analysis, Writing – Original Draft, Visualization; **Qing Liu:** Investigation; **Shanmei Jiang:** Investigation; **Mengyuan Li:** Visualization; **Yue Zhang:** Conceptualization; **Xiaosheng Wang:** Conceptualization, Methodology, Project administration, Funding acquisition, Writing – Review & Editing, Supervision.

## Acknowledgments

We thank China Pharmaceutical University, for it is support and funding.

## Supplemental Content

Supplemental Content 1. PRISMA 2009 Checklist.

Supplemental Content 2. Characteristics of the 52 studies included in the meta-analysis.

Supplemental Content 3. Characteristics of the studies included in the proportional meta-analysis.

Supplemental Content 4. Heterogeneity results from the proportional meta-analysis.

Supplemental Content 5. A summary of the 13 studies included in the network meta-analysis.

Supplemental Content 6. Cochrane Risk of bias analysis (ROB-2) of five randomized trial studies of COVID-19 treatment.

Supplemental Content 7. Subgroup meta-analysis of six treatments to estimate the impact of each treatment on the severity of COVID-19. The forest plots showing the treatment effect on different subgroups (severe versus non-severe, ICU versus non-ICU, death versus survival, and deterioration versus discharge) by the Mantel-Haenszel random-effect model using REVMAN 5 software.

Supplemental Content 8. meta-regression plots showing the impact of chloroquine or hydroxychloroquine and tocilizumab on the severity of COVID-19. The meta-regression plots showing the treatment effect (log (risk ratio)) versus the number of patients who died or stayed in ICU obtained by the mixed-effect model.

Supplemental Content 9. Forest plots showing outlying and influential studies in eight treatments by externally studentized residuals analysis. Each box represents a summary proportion estimated by the leave-one-out study, and the dashed reference line indicates where the original summary proportion lies. The further a box deviates from the reference line; the more pronounced the impact of the corresponding leaving-out study exerts on the original summary proportion.

Supplemental Content 10. Diagnostic plots showing significant and influential studies by analyses of externally studentized residuals, difference in fits values, Cook’s distances, covariance ratios, tau2, and Q-test of leave-one-out heterogeneity, hat values, and weights.

Supplemental Content 11. Funnel plots showing proportional meta-analysis publication bias by testing for the funnel plot asymmetry using the Egger’s regression test. The Z and *P* values are shown.

Supplemental Content 12. Dot plot showing the treatment ranking by the network meta-analysis. The ranking is based on P scores determined by the point estimates and standard errors of the network meta-analysis.

Supplemental Content 13. Comparison-adjusted funnel plot showing the network meta-analysis publication bias by testing for the funnel plot asymmetry using the Egger’s regression test.

## Footnotes

1 CI: confidence interval

2 RR: risk ratio

3 RRT: renal replacement therapy

4 F: F-test

## References

[1] University JH. coronavirus resource center. https://coronavirus.jhu.edu/map.html.

[2] Ksiazek TG, Erdman D, Goldsmith CS, et al. A novel coronavirus associated with severe acute respiratory syndrome. N Engl J Med. 2003;348(20):1953–1966.

[3] Baharoon S, Memish ZA. MERS-CoV as an emerging respiratory illness: A review of prevention methods. Travel Med Infect Dis. 2019:101520.

[4] Rabaan AA, Al-Ahmed SH, Haque S, et al. SARS-CoV-2, SARS-CoV, and MERS-COV: A comparative overview. Infez Med. 2020;28(2):174–184.

[5] *Review Manger Web (RevMan Web)* [computer program]. 2019.

[6] Wang N. How to Conduct a Meta-Analysis of Proportions in R: A Comprehensive Tutorial. 2018.

[7] von Elm E, Altman DG, Egger M, Pocock SJ, Gøtzsche PC, Vandenbroucke JP. The Strengthening the Reporting of Observational Studies in Epidemiology (STROBE) statement: guidelines for reporting observational studies. PLoS Med. 2007;4(10):e296.

[8] Higgins JPT, Altman DG, Gøtzsche PC, et al. The Cochrane Collaboration’s tool for assessing risk of bias in randomised trials. 2011;343:d5928.

[9] Yu M, Liu Y, Xu D, Zhang R, Lan L, Xu H. Prediction of the Development of Pulmonary Fibrosis Using Serial Thin-Section CT and Clinical Features in Patients Discharged after Treatment for COVID-19 Pneumonia. Korean J Radiol. 2020;21(6):746–755.

[10] Xu K, Chen Y, Yuan J, et al. Factors associated with prolonged viral RNA shedding in patients with COVID-19. Clin Infect Dis. 2020.

[11] Wang D, Hu B, Hu C, et al. Clinical Characteristics of 138 Hospitalized Patients With 2019 Novel Coronavirus-Infected Pneumonia in Wuhan, China. JAMA. 2020.

[12] Richardson S, Hirsch JS, Narasimhan M, et al. Presenting Characteristics, Comorbidities, and Outcomes Among 5700 Patients Hospitalized With COVID-19 in the New York City Area. JAMA. 2020.

[13] Ouyang Y, Yin J, Wang W, et al. Down-regulated gene expression spectrum and immune responses changed during the disease progression in COVID-19 patients. Clin Infect Dis. 2020.

[14] Myers LC, Parodi SM, Escobar GJ, Liu VX. Characteristics of Hospitalized Adults With COVID-19 in an Integrated Health Care System in California. JAMA. 2020.

[15] Mo P, Xing Y, Xiao Y, et al. Clinical characteristics of refractory COVID-19 pneumonia in Wuhan, China. Clin Infect Dis. 2020.

[16] Liu Y, Yang Y, Zhang C, et al. Clinical and biochemical indexes from 2019-nCoV infected patients linked to viral loads and lung injury. Sci China Life Sci. 2020;63(3):364–374.

[17] Lian J, Jin X, Hao S, et al. Analysis of Epidemiological and Clinical features in older patients with Corona Virus Disease 2019 (COVID-19) out of Wuhan. Clin Infect Dis. 2020.

[18] Jacobs JP, Stammers AH, St Louis J, et al. Extracorporeal Membrane Oxygenation in the Treatment of Severe Pulmonary and Cardiac Compromise in COVID-19: Experience with 32 patients. ASAIO J. 2020.

[19] Grein J, Ohmagari N, Shin D, et al. Compassionate Use of Remdesivir for Patients with Severe Covid-19. N Engl J Med. 2020.

[20] Bhatraju PK, Ghassemieh BJ, Nichols M, et al. Covid-19 in Critically Ill Patients in the Seattle Region — Case Series. N Engl J Med. 2020.

[21] Huang M, Tang T, Pang P, et al. Treating COVID-19 with Chloroquine. J Mol Cell Biol. 2020;12(4):322–325.

[22] Wang Y, Zhang D, Du G, et al. Remdesivir in adults with severe COVID-19: a randomised, double-blind, placebo-controlled, multicentre trial. The Lancet. 2020.

[23] Tang W, Cao Z, Han M, et al. hydroxychloroquine in patients with mainly mild to moderate coronavirus disease 2019: open label, randomised controlled trial. BMJ. 2020;369:m1849.

[24] Singh S, Chakravarty T, Chen P, et al. Allogeneic cardiosphere-derived cells (CAP-1002) in critically ill COVID-19 patients: compassionate-use case series. Basic Res Cardiol. 2020;115(4):36.

[25] Lian N, Xie H, Lin S, Huang J, Zhao J, Lin Q. Umifenovir treatment is not associated with improved outcomes in patients with coronavirus disease 2019: a retrospective study. Clin Microbiol Infect. 2020.

[26] Hung IF, Lung KC, Tso EY, et al. Triple combination of interferon beta-1b, lopinavir-ritonavir, and ribavirin in the treatment of patients admitted to hospital with COVID-19: an open-label, randomised, phase 2 trial. Lancet. 2020.

[27] Hu K, Guan WJ, Bi Y, et al. Efficacy and Safety of Lianhuaqingwen Capsules, a repurposed Chinese Herb, in Patients with Coronavirus disease 2019: A multicenter, prospective, randomized controlled trial. Phytomedicine. 2020:153242.

[28] Geleris J, Sun Y, Platt J, et al. Observational Study of Hydroxychloroquine in Hospitalized Patients with Covid-19. N Engl J Med. 2020.

[29] Gautret P, Lagier JC, Parola P, et al. Hydroxychloroquine and azithromycin as a treatment of COVID-19: results of an open-label non-randomized clinical trial. Int J Antimicrob Agents. 2020:105949.

[30] Deng L, Li C, Zeng Q, et al. Arbidol combined with LPV/r versus LPV/r alone against Corona Virus Disease 2019: A retrospective cohort study. J Infect. 2020.

[31] Colaneri M, Bogliolo L, Valsecchi P, et al. Tocilizumab for Treatment of Severe COVID-19 Patients: Preliminary Results from SMAtteo COvid19 REgistry (SMACORE). Microorganisms. 2020;8(5).

[32] CHEN Jun LD, LIU Li, LIU Ping,XU Qingnian,XIA Lu,LING Yun,HUANG Dan,SONG Shuli,ZHANG Dandan,QIAN Zhiping,LI Tao,SHEN Yinzhong,LU Hongzhou. A pilot study of hydroxychloroquine in treatment of patients with common coronavirus disease-19 (COVID-19). J Zhejiang Univ (Med Sci). 2020;49(1):0–0.

[33] Zheng S, Fan J, Yu F, et al. Viral load dynamics and disease severity in patients infected with SARS-CoV-2 in Zhejiang province, China, January-March 2020: retrospective cohort study. BMJ. 2020;369:m1443.

[34] Zhao XY, Xu XX, Yin HS, et al. Clinical characteristics of patients with 2019 coronavirus disease in a non-Wuhan area of Hubei Province, China: a retrospective study. BMC Infect Dis. 2020;20(1):311.

[35] Zhang J, Yu M, Tong S, Liu LY, Tang LV. Predictive factors for disease progression in hospitalized patients with coronavirus disease 2019 in Wuhan, China. J Clin Virol. 2020;127:104392.

[36] Yu Y, Xu D, Fu S, et al. Patients with COVID-19 in 19 ICUs in Wuhan, China: a cross-sectional study. Crit Care. 2020;24(1):219.

[37] Yang X, Yu Y, Xu J, et al. Clinical course and outcomes of critically ill patients with SARS-CoV-2 pneumonia in Wuhan, China: a single-centered, retrospective, observational study. Lancet Respir Med. 2020.

[38] Cao B, Wang Y, Wen D, et al. A Trial of Lopinavir-Ritonavir in Adults Hospitalized with Severe Covid-19. N Engl J Med. 2020.

[39] Zhou F, Yu T, Du R, et al. Clinical course and risk factors for mortality of adult inpatients with COVID-19 in Wuhan, China: a retrospective cohort study. Lancet. 2020;395(10229):1054–1062.

[40] Xu XW, Wu XX, Jiang XG, et al. Clinical findings in a group of patients infected with the 2019 novel coronavirus (SARS-Cov-2) outside of Wuhan, China: retrospective case series. BMJ. 2020;368:m606.

[41] Wu J, Liu J, Zhao X, et al. Clinical Characteristics of Imported Cases of COVID-19 in Jiangsu Province: A Multicenter Descriptive Study. Clin Infect Dis. 2020.

[42] Wu C, Chen X, Cai Y, et al. Risk Factors Associated With Acute Respiratory Distress Syndrome and Death in Patients With Coronavirus Disease 2019 Pneumonia in Wuhan, China. JAMA Intern Med. 2020.

[43] Wang Z, Yang B, Li Q, Wen L, Zhang R. Clinical Features of 69 Cases with Coronavirus Disease 2019 in Wuhan, China. Clin Infect Dis. 2020.

[44] Wang D, Yin Y, Hu C, et al. Clinical course and outcome of 107 patients infected with the novel coronavirus, SARS-CoV-2, discharged from two hospitals in Wuhan, China. Crit Care. 2020;24(1):188.

[45] Shekerdemian LS, Mahmood NR, Wolfe KK, et al. Characteristics and Outcomes of Children With Coronavirus Disease 2019 (COVID-19) Infection Admitted to US and Canadian Pediatric Intensive Care Units. JAMA Pediatrics. 2020.

[46] Saleh M, Gabriels J, Chang D, et al. The Effect of Chloroquine, Hydroxychloroquine and Azithromycin on the Corrected QT Interval in Patients with SARS-CoV-2 Infection. Circ Arrhythm Electrophysiol. 2020.

[47] Piva S, Filippini M, Turla F, et al. Clinical presentation and initial management critically ill patients with severe acute respiratory syndrome coronavirus 2 (SARS-CoV-2) infection in Brescia, Italy. J Crit Care. 2020;58:29–33.

[48] Pan L, Mu M, Yang P, et al. Clinical Characteristics of COVID-19 Patients With Digestive Symptoms in Hubei, China: A Descriptive, Cross-Sectional, Multicenter Study. Am J Gastroenterol. 2020;115(5):766–773.

[49] Lo IL, Lio CF, Cheong HH, et al. Evaluation of SARS-CoV-2 RNA shedding in clinical specimens and clinical characteristics of 10 patients with COVID-19 in Macau. Int J Biol Sci. 2020;16(10):1698–1707.

[50] Liu W, Tao Z-W, Wang L, et al. Analysis of factors associated with disease outcomes in hospitalized patients with 2019 novel coronavirus disease. Chin Med J. 2020;133(9):1032–1038.

[51] Liu K, Fang Y-Y, Deng Y, et al. Clinical characteristics of novel coronavirus cases in tertiary hospitals in Hubei Province. Chin Med J. 2020:10.1097/CM1099.0000000000000744.

[52] Jiang Y, He S, Zhang C, et al. Clinical characteristics of 60 discharged cases of 2019 novel coronavirus-infected pneumonia in Taizhou, China. Ann Transl Med. 2020;8(8):547.

[53] Huang C, Wang Y, Li X, et al. Clinical features of patients infected with 2019 novel coronavirus in Wuhan, China. Lancet. 2020;395(10223):497–506.

[54] Hu L, Chen S, Fu Y, et al. Risk Factors Associated with Clinical Outcomes in 323 COVID-19 Hospitalized Patients in Wuhan, China. Clin Infect Dis. 2020.

[55] Hong KS, Lee KH, Chung JH, et al. Clinical Features and Outcomes of 98 Patients Hospitalized with SARS-CoV-2 Infection in Daegu, South Korea: A Brief Descriptive Study. Yonsei Med J. 2020;61(5):431–437.

[56] Chen N, Zhou M, Dong X, et al. Epidemiological and clinical characteristics of 99 cases of 2019 novel coronavirus pneumonia in Wuhan, China: a descriptive study. Lancet. 2020;395(10223):507–513.

[57] Chen H, Guo J, Wang C, et al. Clinical characteristics and intrauterine vertical transmission potential of COVID-19 infection in nine pregnant women: a retrospective review of medical records. The Lancet. 2020;395(10226):809–815.

[58] Cao J, Tu WJ, Cheng W, et al. Clinical Features and Short-term Outcomes of 102 Patients with Corona Virus Disease 2019 in Wuhan, China. Clin Infect Dis. 2020.

[59] Cai J, Xu J, Lin D, et al. A Case Series of children with 2019 novel coronavirus infection: clinical and epidemiological features. Clinical infectious diseases: an official publication of the Infectious Diseases Society of America. 2020:ciaa198.

[60] Beigel JH, Tomashek KM, Dodd LE, et al. Remdesivir for the Treatment of Covid-19 – Preliminary Report. N Engl J Med. 2020.

[61] Chaimani A, Higgins JP, Mavridis D, Spyridonos P, Salanti G. Graphical tools for network meta-analysis in STATA. PLoS One. 2013;8(10):e76654.

[62] Cascella M, Rajnik M, Cuomo A, Dulebohn SC, Di Napoli R. Features, evaluation and treatment coronavirus (COVID-19). In: Statpearls [internet]. StatPearls Publishing; 2020.

[63] Guan WJ, Ni ZY, Hu Y, et al. Clinical Characteristics of Coronavirus Disease 2019 in China. N Engl J Med. 2020;382(18):1708–1720.

[64] Saghazadeh A, Rezaei N. Towards treatment planning of COVID-19: Rationale and hypothesis for the use of multiple immunosuppressive agents: Anti-antibodies, immunoglobulins, and corticosteroids. Int Immunopharmacol. 2020;84:106560.

[65] Cortegiani A, Ippolito M, Ingoglia G, Einav S. Chloroquine for COVID-19: rationale, facts, hopes. Critical Care. 2020;24(1):210.

